# COVID-19 Medical Vulnerability Indicators: A Local Data Model for Equity in Public Health Decision-Making

**DOI:** 10.1101/2020.11.02.20215657

**Authors:** Paul M. Ong, Chhandara Pech, Nataly Rios Gutierrez, Vickie Mays

## Abstract

**Objective:** To develop indicators of vulnerability for coronavirus disease 2019 (covid-19) infection in Los Angeles County (LAC) by race and neighborhood characteristics.

**Design:** Development of indicators that combines pre-existing medical vulnerabilities with social and built-environment data by zip code tabulation areas (ZCTAs).

**Setting:** Neighborhoods in LAC categorized by race/ethnicity ranked into quintiles by relative vulnerability: Non-Hispanic white; Black; Latinx; Cambodians, Hmong and Laotians combined (CHL); and Other Asians.

**Data Sources:** AskCHIS Neighborhood Edition, American Community Survey 2014-2018, and California Department of Parks and Recreation.

**Main Outcome Measures:** 1) Pre-Existing Health Condition, 2) Barriers to Accessing Healthcare, 3) Built Environment Risk, and 4) CDC’s Social Vulnerability.

**Results:** Neighborhoods most vulnerable to COVID-19 are characterized by significant clustering of racial minorities, low income households and unmet medical needs. An overwhelming 73% of Blacks reside in the neighborhoods with the two highest quintiles of pre-existing health conditions, followed by Latinx (70%) and CHL (60%), while 60% of whites reside in low or the lowest vulnerable neighborhoods. For the Barriers to Accessing Healthcare indicator, 40% of Latinx reside in the highest vulnerability places followed by Blacks, CHL and other Asians (29%, 22%, and 16% respectively), compared with only 7% of Whites reside in such neighborhoods. The Built Environment Indicator finds CHL (63%) followed by Latinx (55%) and Blacks (53%) reside in the neighborhoods designated as high or the highest vulnerability compared to 32% of Whites residing in these neighborhoods. The Social Vulnerability Indicator finds 42% of Blacks and Latinx and 38% of CHL residing in neighborhoods of high vulnerability compared with only 8% of Whites residing these neighborhoods.

**Conclusions:** Vulnerability to covid-19 infections differs by neighborhood and racial/ethnic groups. Our vulnerability indicators when utilized in decision-making of re-openings or resource distribution such as testing, vaccine distribution, hotel rooms for quarantine and other covid-19-related resources can provide an equity driven data approach for the most vulnerable.

## Introduction

Los Angeles County, like the rest of the country, is facing a high number of COVID-19 infections and deaths. The county has been affected by the virus at especially high rates, with a total of 289,366 infections and 6,877 deaths as of October 19, 2020^1^. While originally it was thought that all persons had the same level of susceptibility to COVID-19, not all persons nor communities have been equally affected by this disease. Considered the “hotspot” for COVID-19 infections in the state of California, the numbers of COVID-19 infections and deaths in Los Angeles County have not been similarly distributed across neighborhoods. In examining County data, analyses show significant disparities in reported cases and deaths by race and socioeconomic status. These analyses support the notion of inequality in vulnerability in the SARS-CoV-2 (COVID-19) pandemic^2,3,4^.

Understanding how the geographic patterns of social determinants of health and social risk contribute to the medical vulnerability for COVID-19 is useful in helping public health and social service agencies, as well as other stakeholders, to develop and target interventions to the communities at greatest risk for the infectious transmission of COVID-19. This requires developing a comprehensive monitoring system that combines multiple sources of local spatial data to precisely track the temporal and geographic pattern of new cases and to uncover the factors and mechanisms behind the transmission. Such knowledge is critical to slowing and hopefully stopping the pandemic. We are still short of meeting the goal of having a fully operational monitoring system, but there has been progress. The indicators in this brief contribute to that effort, with a focus on identifying vulnerable neighborhoods.

This article draws on recent scientific literature on COVID-19 medical vulnerability, as well as expert methodological input, to better understand factors contributing to the unequal burden of COVID-19 across communities and populations in Los Angeles County. Several studies have identified pre-existing conditions, such as Type 2 diabetes, chronic kidney disease, immunocompromised state and/or severe obesity, that increase risk of COVID-19 infection and complications^5,6,7,8,9,10^. Additionally, limited access to both health protecting equipment and risk and care information can increase the likelihood COVID-19 comorbidities. Furthermore, lack of access to quality healthcare services and social policies, such as access to health insurance, sick days and the ability quarantine in isolation, can further exacerbate the inequality in COVID-19 outcomes^11,12,13,14,15^. Beyond factors directly associated with health, structural factors further increase COVID-19 inequity^16^. Previous research has found that existing spatial inequality is reproduced over time, specifically that urban spatial structures produce and reproduce socioeconomic inequalities^17,18^. The racial discrepancies among COVID-19 outcomes in the U.S. have not only been attributed to COVID-19 comorbidities, but also to the social, economic, and physical factors that provide communities with the capacity to safely practice physical distancing in order to reduce COVID-19 community spread^19^. While much has been made of herd immunity^20,21^ in the absence of a vaccine that can attempt to accomplish that status, far more important at this point is focusing on how to employ data to identify communities whose medical vulnerabilities occur in the context of social and environmental risks and make them highly likely to be exposed, become infected and potentially suffer significant and costly morbidity and mortality outcomes^10, 22^.

The purpose of this project is to develop multiple indicators that point to probable communities (geographic places defined by the Census Bureau’s Zip Code Tabulation Areas) and populations at risk in Los Angeles County with high probability of COVID-19 infection and death across different dimensions. To achieve this, we developed a medical vulnerability index of four indicators: pre-existing health vulnerability, *barriers to accessing healthcare, built environment risk*, and *social vulnerability*. We hope that these indicators will be helpful for policy makers, local jurisdictions, foundations, and community organizations to identify areas with a high need of resources to protect against a potential second wave of COVID-19 infections. Additionally, we hope this information will help determine equitable distribution of resources to stem new infections such as testing and vaccination, if available, in order to create more equitable and healthy neighborhoods, especially for medically vulnerable populations that live in those neighborhoods. It is our hope that this brief can help guide local communities in how to aggregate their own local data to help drive initiatives that can respond to COVID-19 with greater equity. Local regions will vary in the ability to harness local data and even in our instance there were many discussions devoted to wanting data sources that were either not available, or would be time or cost prohibitive to develop.

We present a detailed description of our methodology. The description includes information about our data sources as well as information on how we developed each of our four indicators. We provide data visualizations as part of our analyses to indicate geographic patterns by neighborhoods. First, we include multiple maps in order to illustrate vulnerability across our four indicators. Second, we present two different analytical components: 1) an analysis showcasing the racial distribution across each of the four indicators; and 2) an analysis showcasing the average vulnerability scores by majority ethnoracial groups. Last is our discussion and conclusion. Our goal is to assess how our indicators work in illustrating race and place based differences in vulnerability to Covid-19 infections. Our overall goal is to demonstrate how local data can be used to locate, illustrate differences based on not just medical vulnerabilities but social risk which can indicate racial disparities in Covid-19. As local areas must contend with resource allocations, ethical methods for testing and should it be developed vaccine distribution local models of medical vulnerability that attend to social determinants and social risk factors will be critical.

## Data and Methodology

The project focuses on identifying neighborhoods in Los Angeles County that may be at an elevated risk of exposure and positive infection for contracting COVID-19. To achieve this, we developed four different indicators of medical vulnerability: 1) Pre-Existing Health Condition, 2) Barriers to Accessing Healthcare, 3) Built Environment Risk, and 4) Social Vulnerability. Selections of the components used to construct the first three indicators were guided by existing research literature and input from health experts. The fourth indicator, the Social Vulnerability index, mirrors the Center for Disease Control’s (CDC) 2018 Social Vulnerability Index^23^ but constructed for a different geographic unit of analysis then what is made available by the CDC.

### Geographic Unit of Analysis

The basic geographic unit of analysis for this report is the Zip Code Tabulation Area (ZCTA). We utilize the ZCTA because our main source of population level health data, the California Health Interview Survey was only available to use in ZCTA format. The ZCTA is defined by the Bureau of the Census (BOC) as “generalized area representations of United States Postal Service (USPS) ZIP Code service areas”^24^. Zip Codes created by USPS for mail delivery purposes are constantly changing. ZCTAs do not represent actual Zip Codes per se but are made by the Bureau of Census to approximate Zip Codes and their boundaries are defined every 10 years with the Decennial Enumeration. Through ZCTAs, the BOC is able to provide Census related data (e.g. demographic, socioeconomic, housing characteristics) to a geography that closely mirrors USPS Zip Codes.

In some cases, there are occurrences where some of the data used to construct the indicators are reported in a different geography other than ZCTAs. When there are incidents of this, we use a geographic crosswalk to allocate the information into the ZCTA. The geographic crosswalk comes from the Missouri Census Data Center’s (Geocorr 2018 edition). For example, census tracts do not fall completely within ZCTA boundaries. As such, a crosswalk is needed to allocate tract level data into ZCTAs. The spatial assignments take into account differences in population across tracts and therefore a population weighted approach is used to assign tracts to ZCTAs. Below, we indicate which variables are reported in a different geography other than ZCTA and therefore require using the geographic crosswalk to assign to ZCTAs.

For the purpose of this report, ZCTAs are used to represent neighborhoods and the two terms are used interchangeably.

### Data Sources

Three major data sources are used to construct the four indicators of medical vulnerability. These datasets are described below.

#### AskCHIS Neighborhood Edition^25^

The AskCHIS Neighborhood Edition (AskCHIS NE) database provides health estimates for California down to the zip code tabulation area.^4^ The information for AskCHIS NE comes from the California Health Interview Survey (CHIS). CHIS is conducted by the UCLA Center for Health Policy Research (CHPR) in collaboration with the California Department of Public Health and the Department of Health Care Services. CHIS is the largest state health survey in the nation. Conducted since 2001, CHIS surveys adults, adolescents and children sampled from every county in California. CHIS collects extensive information for all age groups on sociodemographic, health status, health conditions, health-related behaviors, health insurance coverage, access to health care services, and other health-related issues. The AskCHIS NE data used for this project represents data collected for 2015-2016 (hereinafter referred to as 2016), which is the most current data available from AskCHIS NE. The data was purchased directly from the CHPR and is not publicly available for download.

#### American Community Survey^26^

The American Community Survey (ACS) is an ongoing survey conducted by the U.S. Census Bureau to collect housing, demographic, social and economic information. The data utilized in this project comes from the 2014-2018 five-year ACS estimates. For small geographies, including ZCTAs, statistics from the ACS are only reported in the five-year average dataset. Small geographic areas reported by the BOC are defined as any area with less than 65,000 persons. Each annual ACS survey represents a sample of about 2.0-2.5% of households and individuals, with the five-year ACS representing roughly 12.5%.

#### California Department of Parks and Recreation^27^

The California Department of Parks and Recreation (DPR) provide data on the availability of parks and open space in California. For this project, we included the DPR’s data on park acres per 1,000 residents^27^, which is reported at the census tract level. Because of the difference in reporting geography, we allocated DPR’s census tract data into ZCTAs using a geographic crosswalk from census tracts to ZCTAs, weighted by the population of each tract.

### Construction of the Indicators

#### Pre-existing Health Vulnerability

- The Pre-existing Health Vulnerability indicator is meant to capture risks of COVID-19 infection and death due to pre-existing health conditions that have been identified in scientific journals and through input from health experts consulted for this project. This indicator is comprised of six different variables, all of which are derived from the AskCHIS NE 2016 database, and which are described below:
- Diabetes, defined as any adult respondent over the age of 18 ever diagnosed with diabetes by a doctor. Diabetes has been identified in the literature as a pre-existing condition increasing risk of COVID-19 infection or death^5,7,9^.
- Obesity, defined as any adult respondent over the age of 18 with a body mass index (BMI) of 30.0 or above. Obesity has also been identified as a pre-existing condition that increases likelihood of COVID-19 complications^6,7,8,9^.
- Heart Disease, defined as any adult respondent over the age of 18 ever diagnosed with heart disease by a doctor. We included heart disease as a dimension in lieu of specific data on hypertension which has been identified as one of the most common comorbidities related to increased COVID-19 risk^5,7,9^.
- Health Status, defined has any adult respondents ages 18-64 with fair or poor health. We included health status as a measure of fair or poor health as a substitute for other pre-existing health conditions absent from our data sources.^i^
- Mental Health, defined as any adult respondent over the age of 18 who reported serious psychological distress in the past 12 months, constructed using the Kessler 6 series (K6 greater or equal to 13). People with severe mental health tend to have higher levels of pre-existing conditions, such as Type 2 diabetes and heart disease, than the general population^28^.
- Food Insecurity, defined as any adult respondent over the age of 18 with income less than 200% below the federal poverty line who self-identified their ability to afford enough food^29^. We included this variable as a measure of poor nutrition. Poor nutrition is a leading factor in contributing to widespread instances of diabetes and obesity across the world.^30^

#### Barriers to Accessing Health Care

The indicator on Barriers to Accessing Health Care is meant to capture barriers that increase difficulty in accessing COVID-19 and other general healthcare services. This indicator is composed of five different variables, all derived from the 2014-18 5-year ACS data:

- Non-U.S. Citizens, defined as the share of immigrants who are not U.S. citizens. We include non-U.S. citizens because this population often faces cultural and legal barriers to accessing healthcare. Most of this group are from non-Western countries, and may risk being labeled a “public charge,” which could potentially jeopardize their immigrant status. A disproportionate number also have limited-English ability, and lack of health insurance, which are factors included separately in the indicator.
- English Language Barrier; defined as the share of the population aged 5 years of older that speak English “less than well”. Language barriers can often prevent people from accessing important information in a timely manner. Many organizations lack the resources necessary to provide documents in multiple-languages. Even if organizations have enough resources, it takes time to provide all the necessary information in languages that are accessible to different communities. This delay in translation applies to information regarding COVID-19 risk and prevention.
- Lack of Broadband Access, defined as the share of households with a computer but without broadband internet access. Lack of broadband hinders access to important information distributed by local and federal health agencies regarding COVID-19, such as where to find the closest COVID-19 testing center.
- Lack of Health Insurance, defined as the share of individuals without health insurance. Despite the fact that the Coronavirus Aid, Relief, and Economic Security Act (CARES Act) reimburses the medical cost of those with COVID-19, lack of healthcare insurance may cause delays in accessing preventive care and seeking other health related benefits. Moreover, many may not know about this provision in the CARES Act.
- Vehicles per Person, defined as the inverted ratio of vehicles available per person. We inverted the ratio to indicate a higher level of vulnerability for households that have fewer cars per person. Depending on the number of cars available per person, having the availability to use a car for medical purposes might not be an option given that there might be other priorities such as getting to work or school that prevents people in the household from getting the healthcare they need. Moreover, some COVID-19 testing sites in Los Angeles require that people arrive in a motorized vehicle. For example, Dodger Stadium serves as LA County’s largest coronavirus drive-thru testing site.

#### Built Environment Risk

The built environment risk indicator is meant to identify areas at higher risk of COVID-19 infection due to lack of adequate space available to adhere to shelter-in-place mandates and other precautions that aim to limit the spread of COVID-19. The indicator is composed of four variables:

- Population density, operationally defined as the total number of persons divided by the ZCTA’s land area in square miles. Counts of the population are derived from the 2014-18 5-year ACS. Places that are densely populated increases the chances of encountering people, which limits the ability to maintain social distancing guidelines and increases the likelihood of encountering a COVID-19 carrier.
- Building structure density, operationally defined as housing structures with 10 or more units divided by the total housing stock (i.e. as a share of all housing units in the ZCTA). Similar to population density, building density also increases chances of encountering people which limits social distance guidelines and increases likelihood of encountering a COVID-19 carrier. We focus on 10 or more units because as opposed to including all multi-units (e.g. duplexes, triplexes), structures with 10 or more units are more likely to increase one odds of encountering people in common areas (e.g. lobby, hallways, mailrooms) and therefore increases the risk and COVID-19 contagion.
- In-unit housing crowding, operationally defined as having 1.01 or more persons per room. In-unit crowding can increase a person’s risk to COVID-19 infection. If someone in a household becomes infected with COVID-19 and there is not a room for them to quarantine in, the rest of the household has a higher risk of contracting the disease. A study recently found that areas with the highest number of COVID-19 cases faced three-times the level of overcrowding than areas with the lowest number of COVID-19 cases^31^.
- Availability of parks and open space per 1,000 residents. Areas with more parks and open space enable individuals to more easily keep physically and mentally fit through outdoor exercise or activity.

#### Social Vulnerability Index

The social vulnerability indicator is a close replica of the 2018 Social Vulnerability Index (SVI), which was created by the Center for Disease Control and Prevention (CDC) to identify vulnerable areas in need of preparation and response to hazardous events or natural disasters. The SVI indicator addresses the level to which community experiences different social conditions, such as unemployment and that might affect its ability to prepare and respond to hazardous events, such as the COVID-19 pandemic. The CDC’s SVI is only available at the census tract level. We utilize the same data source used by the CDC to construct their SVI, 2014-18 5-year ACS, and adopted the same methodological approach to construct the SVI for ZCTAs. A total of 15 variables, organized into four dimensions, are used to construct the SVI for ZCTAs in Los Angeles County. Each component of the SVI is described below.

#### Socioeconomic Status

- Persons Below Poverty, defined as the share of persons with income below the federal poverty line;
- Unemployed, defined as the share of civilian labor force population (ages 16 and up) who are unemployed;
- Per capita Income; is a measure of the amount of income earned per person
- No High School Diploma, defined as share of persons age 25 and older with no high school diploma;

#### Household Composition

- Persons Aged 65 or Older as a share of the total population;
- Persons Aged 17 or Younger as a share of the total population;
- Civilian noninstitutionalized population with a disability; defined as any individuals age 5 years or older with a disability;
- Single-Parent Households with children under 18 as a share of total households;

#### Minority Status and Language

- Racial Minority Population, defined as the share of the population who are not non-Hispanic white (e.g. total population minus non-Hispanic white);
- Speaks English “Less than Well”; defined as the share of the population aged 5 years of older that speak English “less than well”;

#### Housing Type and Transportation

- Multi-Unit Structure, defined as the share of housing structures with 10 housing units or more;
- Mobile Homes, defined as the share of mobile homes;
- In-unit Housing Crowding, defined as having 1.01 or more persons per room;
- No Vehicle Households, define as households with no vehicles available as a share of all households;
- Group Quarters, defined as share of persons in institutionalized group quarters.

### Ranking Methodology

The project adopts a ranking approach to construct each of the four composite indicators. Only ZCTAs within Los Angeles County are included in this process^ii^. We adopted the ranking approach utilized by the CDC to construct their Social Vulnerability Index.^iii^ The advantage of utilizing this approach is consistency in method in constructing the project’s four indicators.

For each of the four composite indicators, we first rank each of the key underlying variables used to construct each index. Using the ranking procedure in SAS, statistical software, we rank each individual variable, with ranking values ranging from 0 to 99, which represent percentile rankings. After ranking each underlying variable, we then summed the percentiles ranking, and conducted a second wave of ranking using the summed percentiles. This in the end represents the actual composite score.

In the case of the Social Vulnerability index which is composed of multiple variables, organized by four key dimensions, three rankings are involved: first, ranking each variable in each dimension, ranking of the sum of the key variables, and lastly a ranking of a sum of all of the variables and dimensions combined.

It is important to note that there are cases where a ZCTA may not have values or data across all underlying variables. In these cases, the ZCTA was excluded. In other words, a ZCTA must have a reported value (no missing data) for each of the underlying variables that are needed to construct the index. If there is missing data for one of the components, then no index is calculated for the ZCTA.

For analytical purposes and for mapping, we rank indicators into quintiles, from lowest to highest vulnerability. Each quintile group contains roughly 56 ZCTAs. Maps for each of the four vulnerability indicators are displayed in the next section.

## Results

### Maps

The following maps display neighborhoods in Los Angeles County by their level of vulnerability across each of the four indicators: *pre-existing conditions, barriers to accessing healthcare, build environment risk*, and *social vulnerability*. ZCTAs are grouped into quintile categories (five groups) based on their composite scores for each of the four indicators, ranging from lowest to highest vulnerability. Each quintile group contains roughly 20% of all ZCTAs within Los Angeles County.

The brown areas represent neighborhoods that are vulnerable, with darker brown denoting the greatest vulnerability. The green areas represent neighborhoods that are less vulnerable, with the darker green denoting the lowest vulnerability. We also provide a map, displayed in Figure 1, using regions defined by the Los Angeles Times to provide as visual aid for understanding the layout of neighborhoods across Los Angeles^iv^.

**Figure 1.**
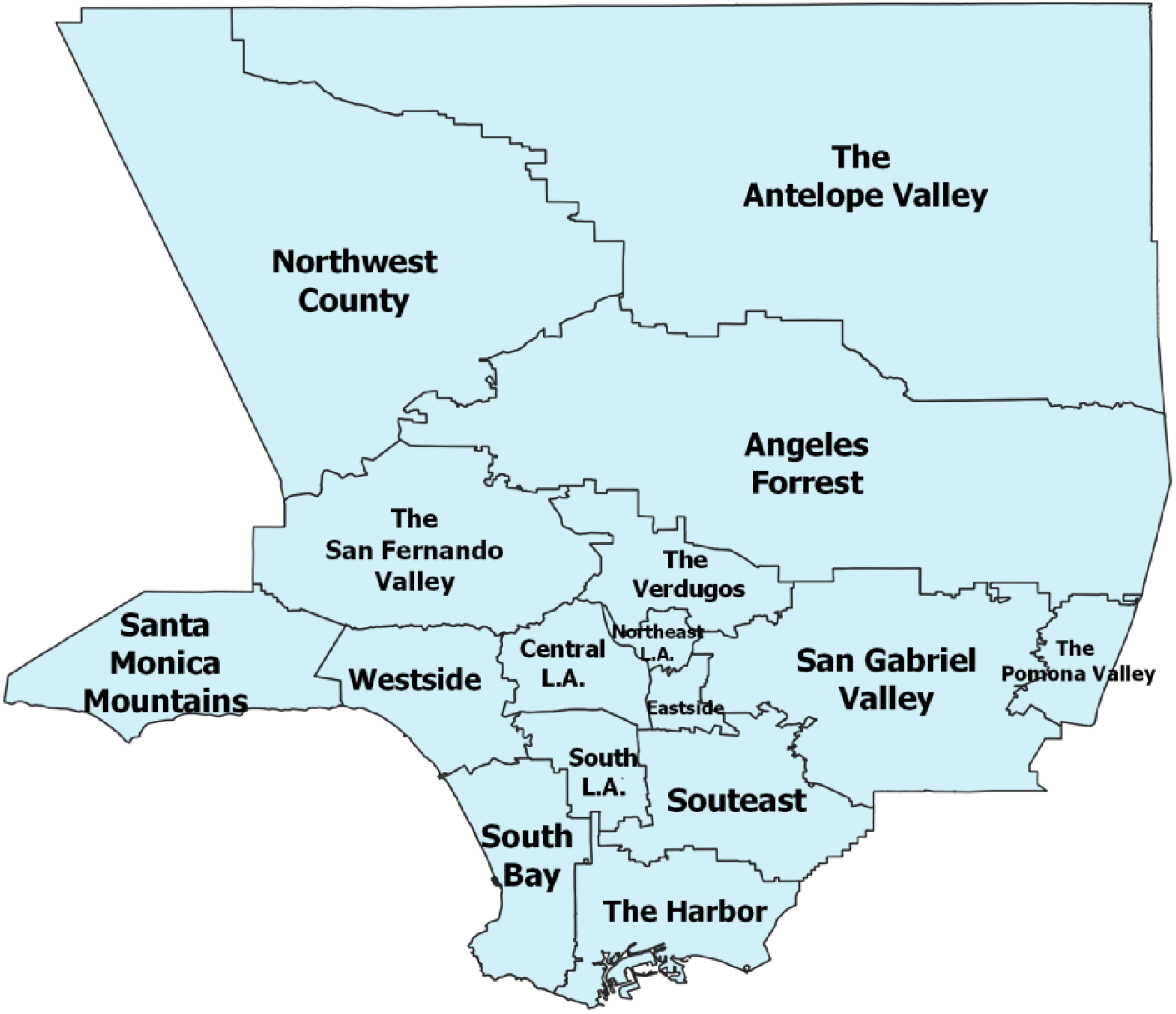
Los Angeles County Regions.

Overall, we find that neighborhoods located in areas around South Los Angeles and the eastern sides of the San Fernando Valley are within the highest vulnerability quintile across all indicators. These areas have a disproportionately high number of low-income households and people of color. In contrast, communities located along the coast and the Northwest County is within the lowest vulnerability across all four indicators. The residents in these neighborhoods are disproportionately non-Hispanic White and high-income.

In the *Pre-existing Health Vulnerability* map, Figure 2, neighborhoods in the high or highest vulnerability quintiles include those in the Antelope Valley, the San Fernando Valley, Boyle Heights and East Los Angeles, and neighborhoods in South Los Angeles. In contrast, the least vulnerable neighborhoods are located in Northwest County as well as across all coastal neighborhoods.

**Figure 2.**
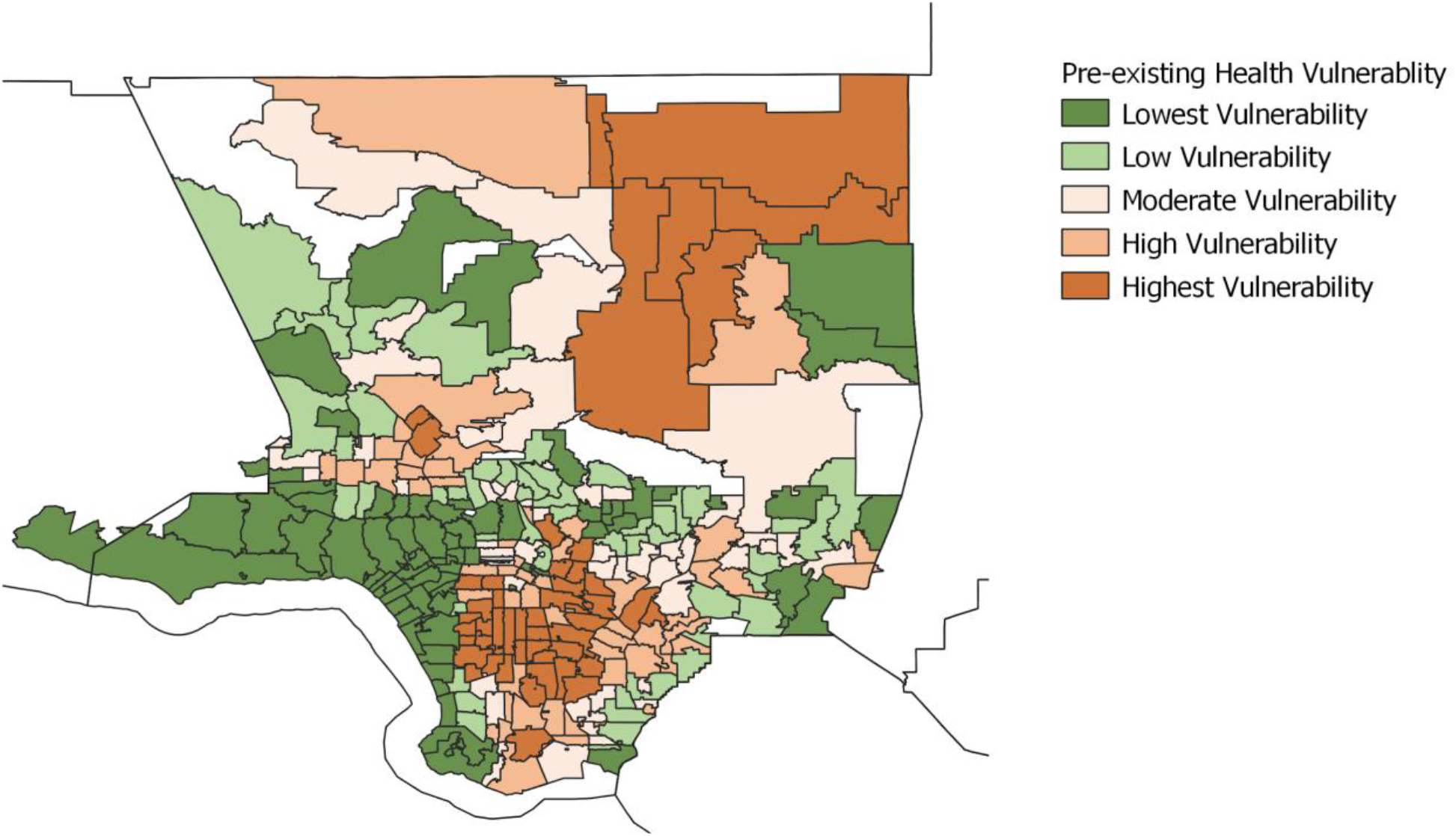
Pre-Existing Health Vulnerability, Los Angeles County. **Figure 2** demonstrates the pre-existing health vulnerability in Los Angeles County, which captures the risks of COVID-19 infection and death as a result of pre-existing health conditions. These conditions include diabetes, obesity, heart disease, health status, mental health, and food insecurity.

Figure 3 displays the map for the *Barriers to Healthcare* indicator. Neighborhoods that are in the high or highest vulnerability are located mainly in Central Los Angeles, South and Southeast L.A., as well as pockets of neighborhoods in the San Fernando and San Gabriel Valley. The neighborhoods falling within the lowest vulnerability are again located along the coast and in Northwest County.

**Figure 3.**
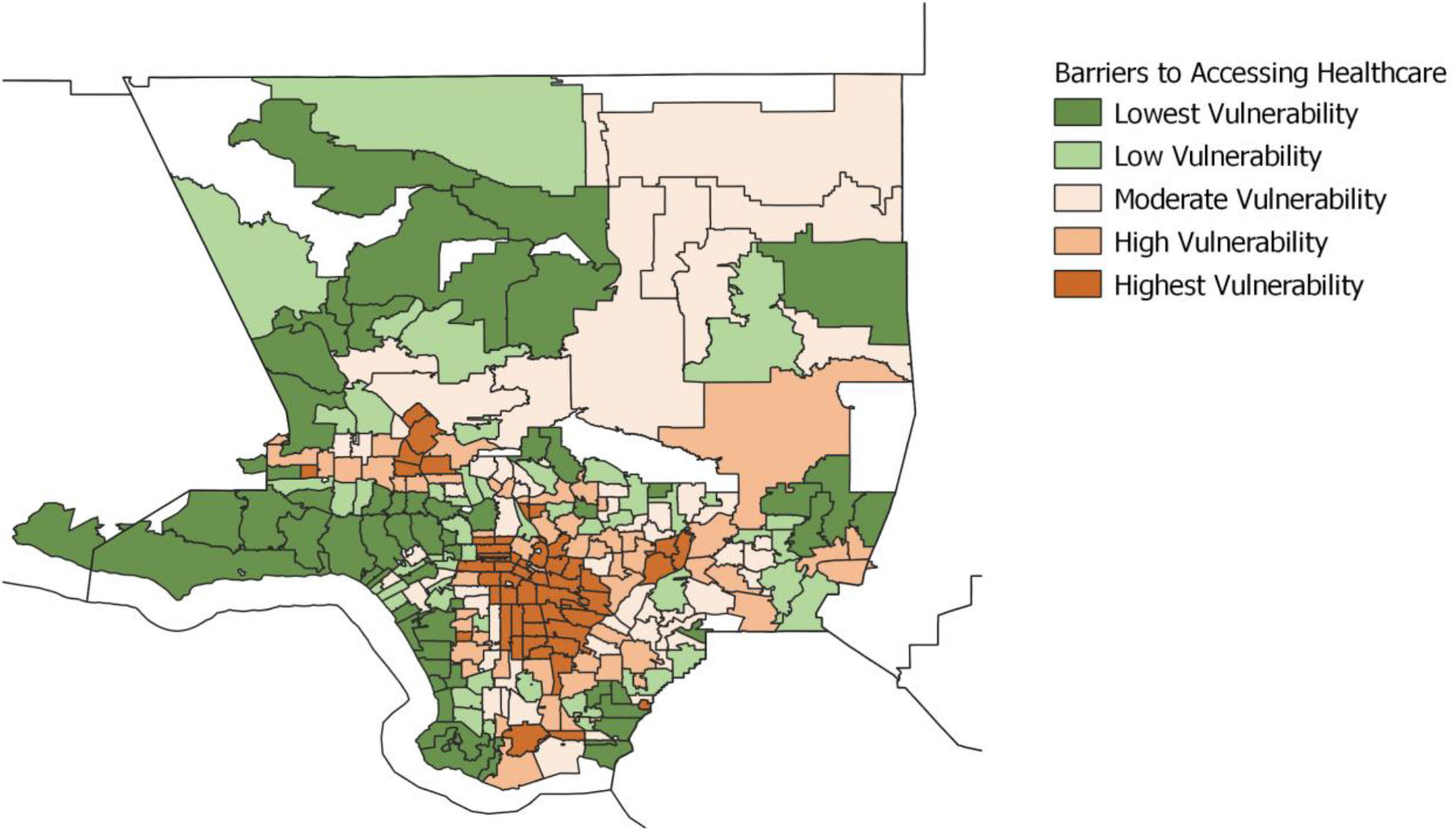
Barriers to Accessing Healthcare, Los Angeles County. **Figure 3** illustrates the vulnerability due to barriers to accessing health care, which captures barriers that increase difficulty in accessing COVID-19 and other general healthcare services. The five variables that make up this indicator include non-U.S. citizen status, English language barrier, lack of broadband access, lack of health insurance, and vehicles per person.

For the *Built Environment Risk* indicator, Figure 4, regions in the high or highest vulnerability quintile are again concentrated in Central L.A., Eastside L.A., South L.A. and San Fernando Valley. In contrast, communities within the lowest quintiles expand around the city’s outskirts and include neighborhoods in Northwest County, the Antelope Valley, and the Santa Monica mountains.

**Figure 4.**
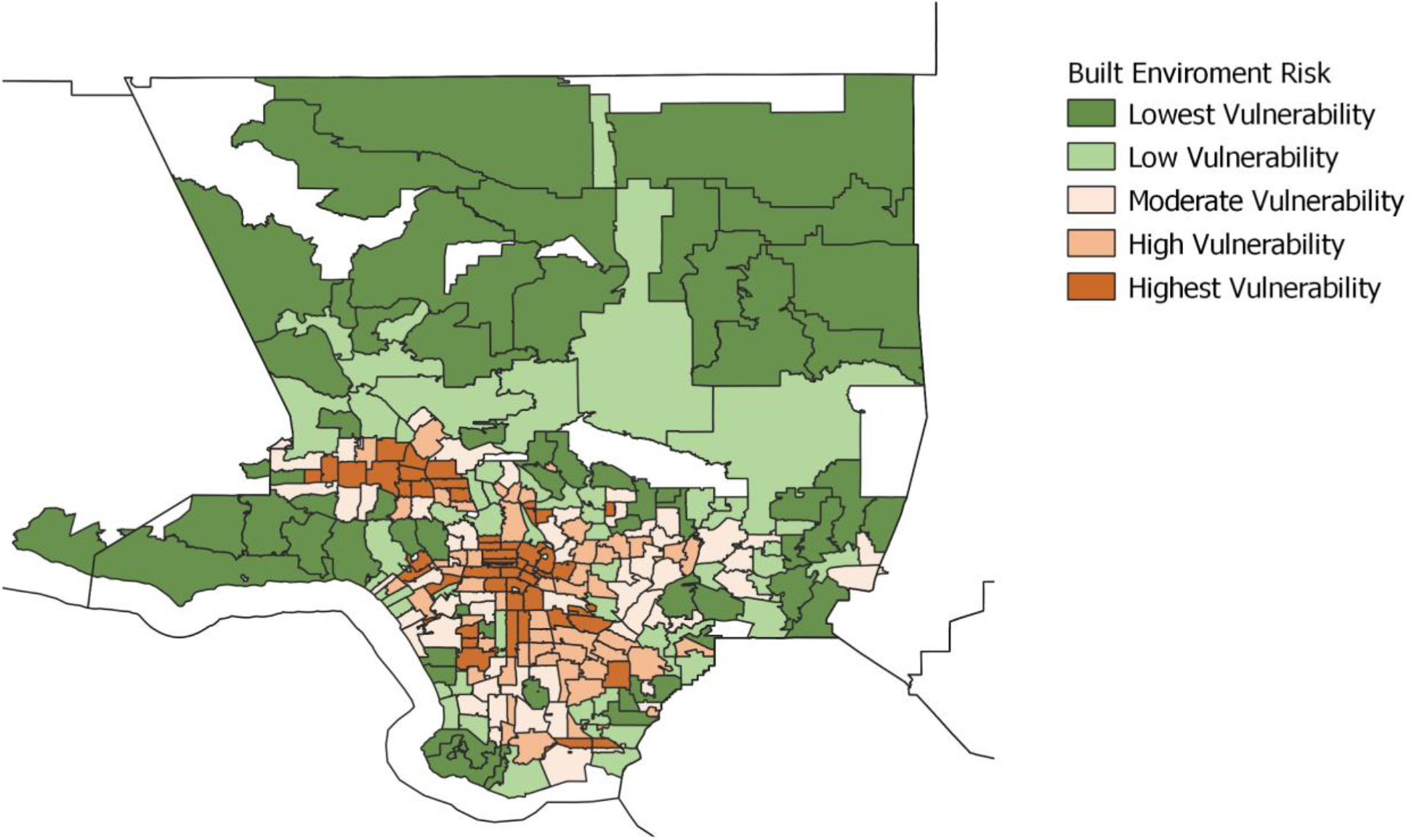
Built Environment Risk, Los Angeles County. **Figure 4** depicts the built environment risk indicator, which identifies areas at higher risk for COVID-19 infection due to lack of adequate space available to follow shelter-in-place mandates and other precautions necessary to limit the spread of COVID-19. Four variables define this indicator: population density, building structure density, in-unit housing crowding, and availability of parks and open space per 1,000 residents.

Finally, neighborhoods in the high and highest vulnerability quintiles for the *Social Vulnerability Index*, Figure 5, are located in the Antelope Valley, the San Fernando Valley, Northeast L.A., Eastside, South and Southeast L.A., and the Harbor. The lowest SVI vulnerability neighborhoods are the coastal communities and in Northwest County.

**Figure 5.**
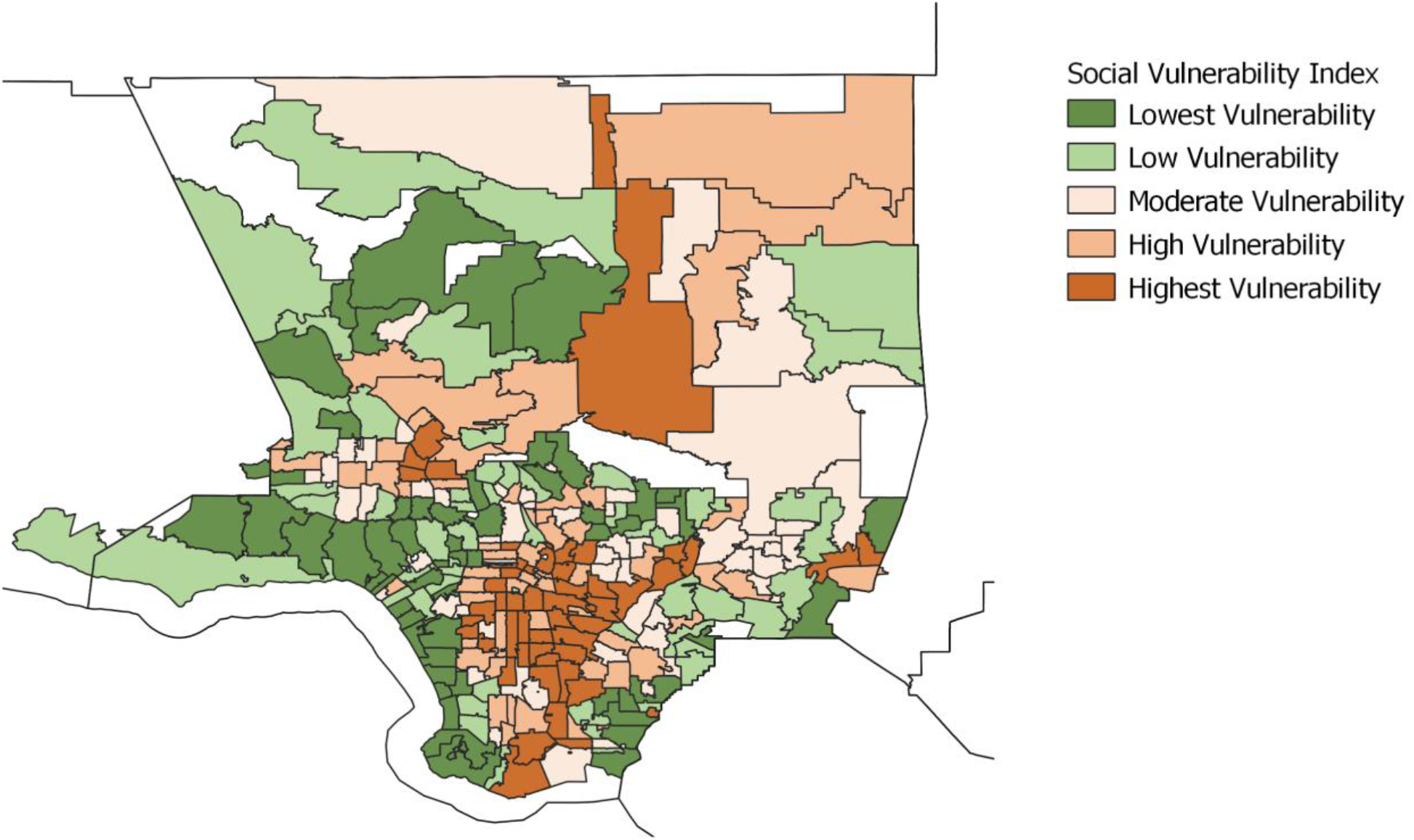
Social Vulnerability Index, Los Angeles County. **Figure 5** captures the social vulnerability index, which identifies vulnerable areas in need of preparation and response to hazardous events or natural disasters. The four components of this index include socioeconomic status, household composition, minority status and language, and housing type and transportation.

### Data Analysis Results for Indicators

For this report’s analytical component, we investigated the racial distribution of vulnerability across each of the four indicators. We further explore racial parity by examining the level of vulnerability across the four indicators based on the majority race for each ZCTA. Majority race is defined as having an ethnoracial composition greater or equal to 50% for a particular ZCTA (e.g. if the population of Blacks in a ZCTA is 50% or more then the ZCTA is designated as a “Majority Black” neighborhood). (Appendix A includes additional analysis of variations by neighborhood socio-demographic characteristics)

#### Ethnoracial Distribution of Vulnerability

The purpose of this analysis is to understand the distribution of different ethnoracial groups across each vulnerability indicator. In each of the figures below, each bar represents a given racial group, and segments denote the percent population within a given vulnerability quintile for each indicator. The data is ranked from the lowest to the highest vulnerability. Brown indicates a high vulnerability, with a darker brown indicating a higher vulnerability level. Green represents lower vulnerability, with a darker green indicating a lower level of vulnerability.

We included five ethnoracial groups: Non-Hispanic white (NH White); Black; Latinx; Cambodians, Hmongs and Laotians combined (CHL Asians); and Other Asians. Asian Americans are a heterogeneous ethnoracial group and also diverse with respect to socioeconomic outcomes. We separate out Cambodians, Hmongs, and Laotians from other Asians since these groups tend to be among the most economically disadvantaged within the Asian American subgroups in Los Angeles County.

Figure 6, illustrates the uneven distribution of *Pre-Existing Health Vulnerability* across racial groups. Black individuals carry the highest burden of pre-existing health conditions. Nearly 3/4th (73%) of the African American population in the county reside in neighborhoods designated as either high or highest vulnerability as measured by the pre-existing health condition index. In contrast, only 8% of the Black population resides in neighborhoods with the lowest level of vulnerability. Similarly, a high share of the Latinx and CHL Asian populations also reside in the most vulnerable neighborhoods. About 70% of the Latinx and 60% of the CHL Asian population reside in neighborhoods with either high or highest levels of vulnerability. In comparison, only about 5% and 4% of their respective population live in neighborhoods with the lowest level of vulnerability. Conversely, neighborhoods in the lowest vulnerable category contain a higher percent of non-Hispanic whites when compared with the highest vulnerable category. Close to 60% of the NH white population in the county reside in either the low or lowest vulnerable neighborhoods.

**Figure 6.**
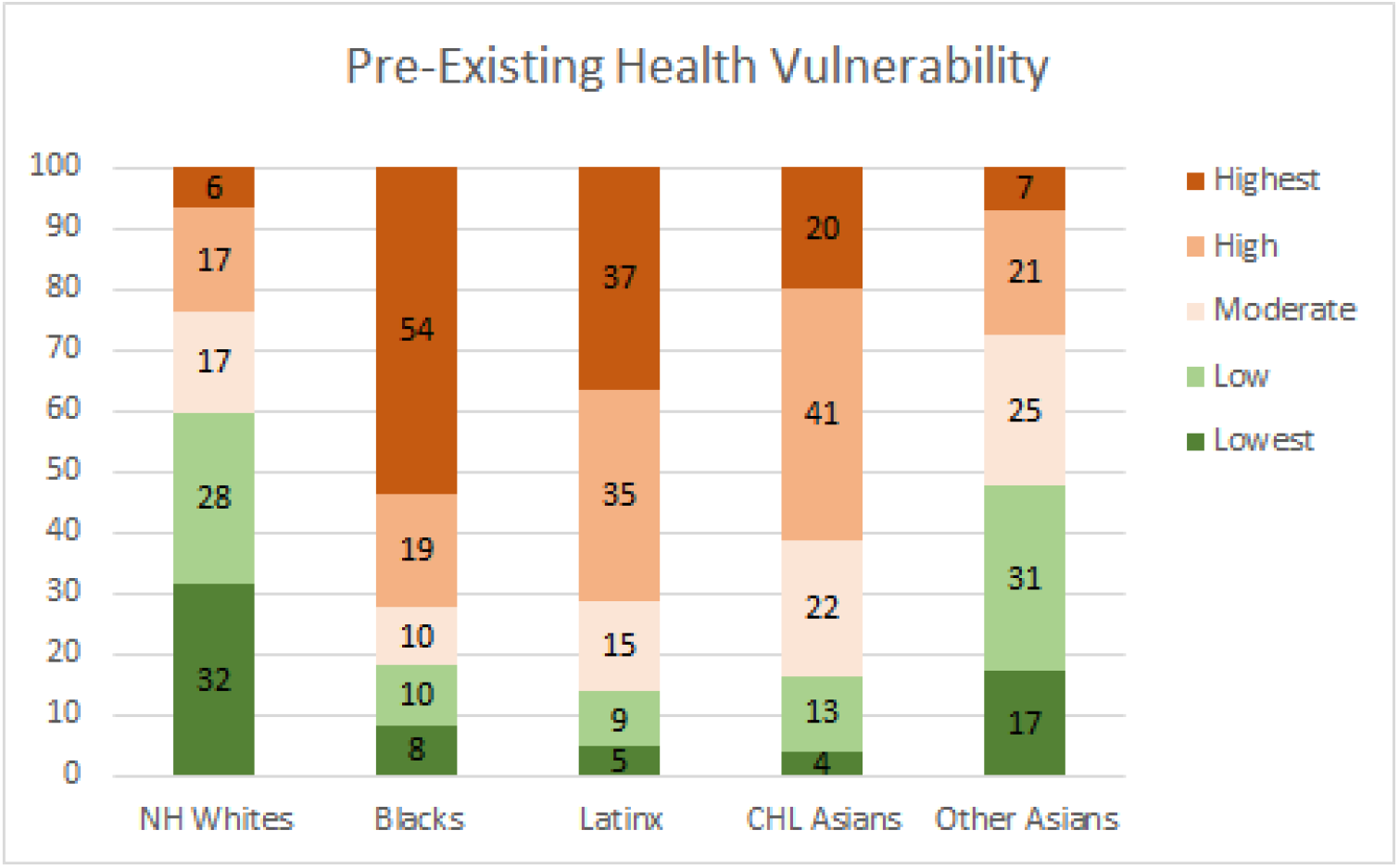
Ethnoracial Distribution by Pre-Existing Health Vulnerability

Similarly, in Figure 7, we find that people of color are more likely to reside in neighborhoods with more *Barriers to Accessing Healthcare*. Forty percent of the Latinx population resides in neighborhoods with the highest level of vulnerability, while only 5% of the population resides in neighborhoods with the lowest level of vulnerability. Black, CHL Asian, and Other Asians are also disproportionately located in neighborhoods with the highest level of vulnerability (29%, 22%, and 16% respectively). In contrast, 7% of the NH White population resides in neighborhoods with the highest level of vulnerability, while 31% of whites reside in neighborhoods with the lowest level of vulnerability.

**Figure 7.**
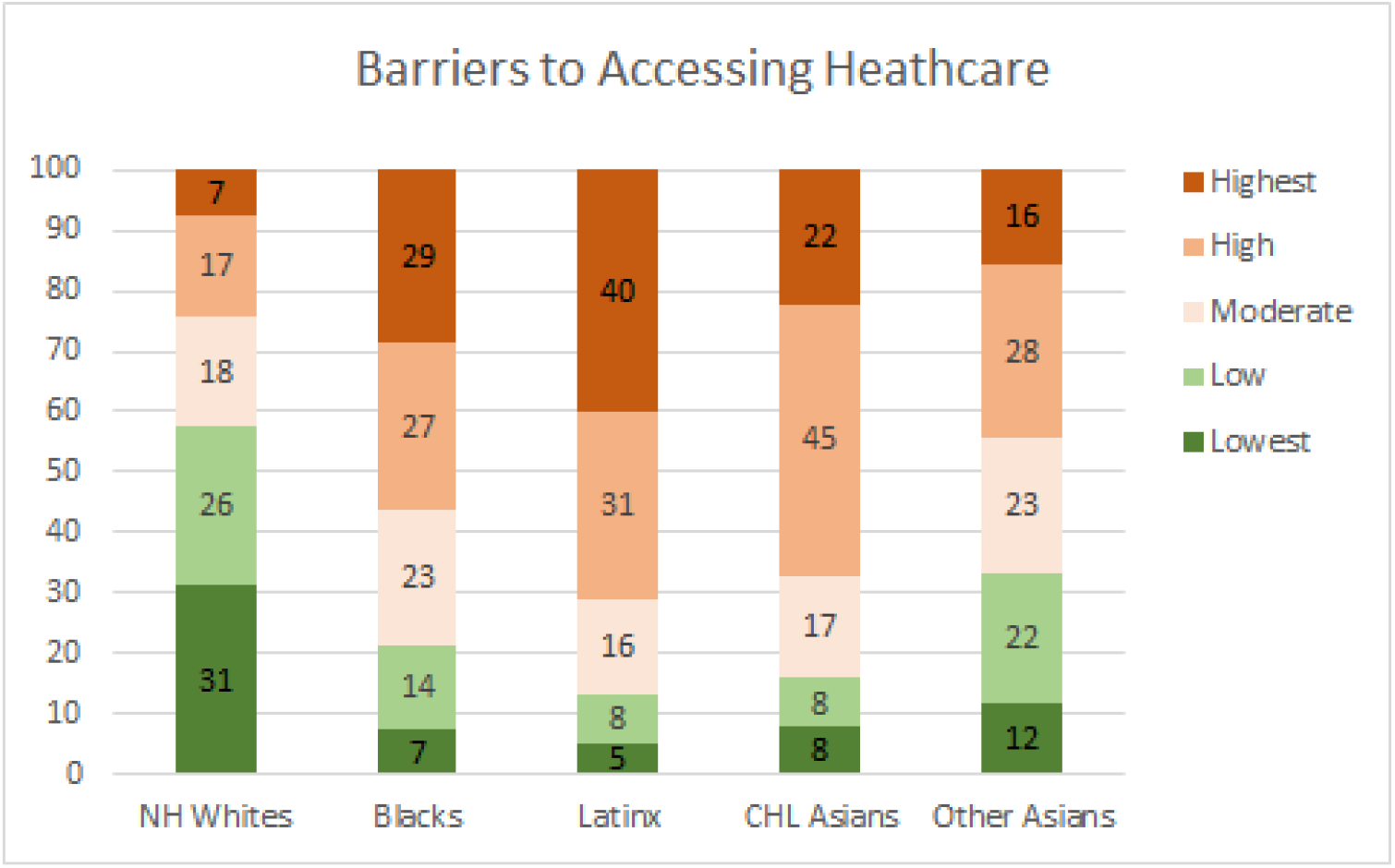
Ethnoracial Distribution by Neighborhood Barriers to Accessing Healthcare

In Figure 8, we observe that in the *Built-Environment Risk indicator*, Black, Latinx and CHL Asian populations are heavily concentrated in neighborhoods designated as either high or highest levels of vulnerability (53%, 55%, 63% respectively). In contrast, about one third (32%) of the county’s NH White population reside in high or highest vulnerability neighborhoods as it relates to the built environment, while almost half (49%) reside in neighborhoods with the lowest level of vulnerability.

**Figure 8.**
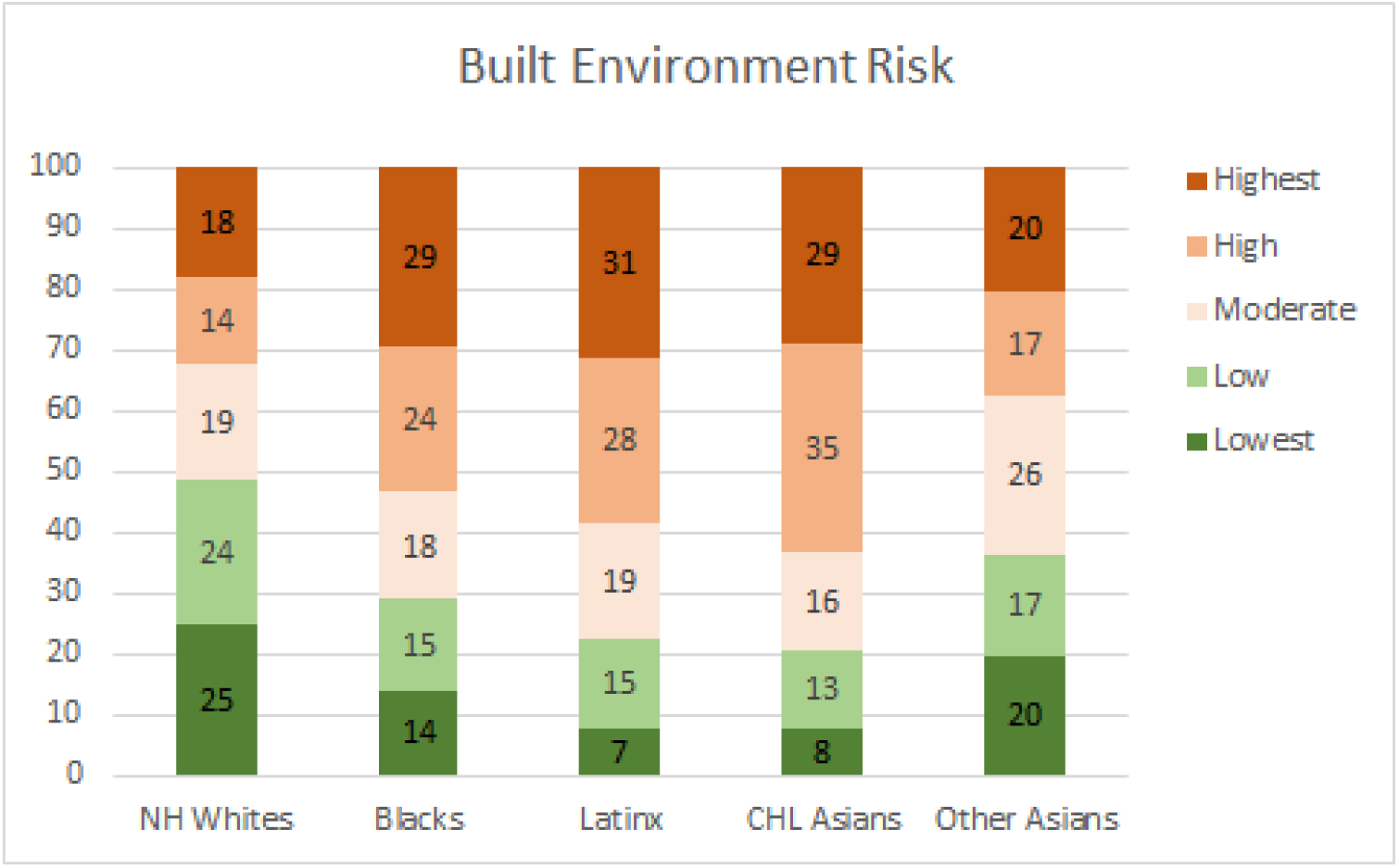
Ethnoracial Distribution by Neighborhood Built Environment Vulnerability

Lastly, Figure 9, displays the five ethnoracial groups’ distribution by the neighborhoods Social Vulnerability Index. This process is partly tautological because the Social Vulnerability Index includes the shared minority population; nonetheless, the graph does demonstrate a disproportionate distribution of vulnerability across different racial and ethnic communities. The Black, Hispanic, and CHL Asian populations reside in neighborhoods with the highest level of social vulnerability. About 42% of the Black population resides in neighborhoods with the highest vulnerability, while only 7% reside in neighborhoods with the lowest vulnerability. Similarly, about 42% of the Latinx population resides in neighborhoods with the highest vulnerability, while only 5% reside in neighborhoods with the lowest vulnerability. Likewise, about 38% of the CHL Asian population resides in neighborhoods with the highest vulnerability, while only 9% reside in neighborhoods with the lowest vulnerability. Again, the NH White population resides mainly in neighborhoods with the lowest level of vulnerability (31%) as measured by the SVI, while only a small share of their population (8%) resides in neighborhoods with the highest level of vulnerability.

**Figure 9.**
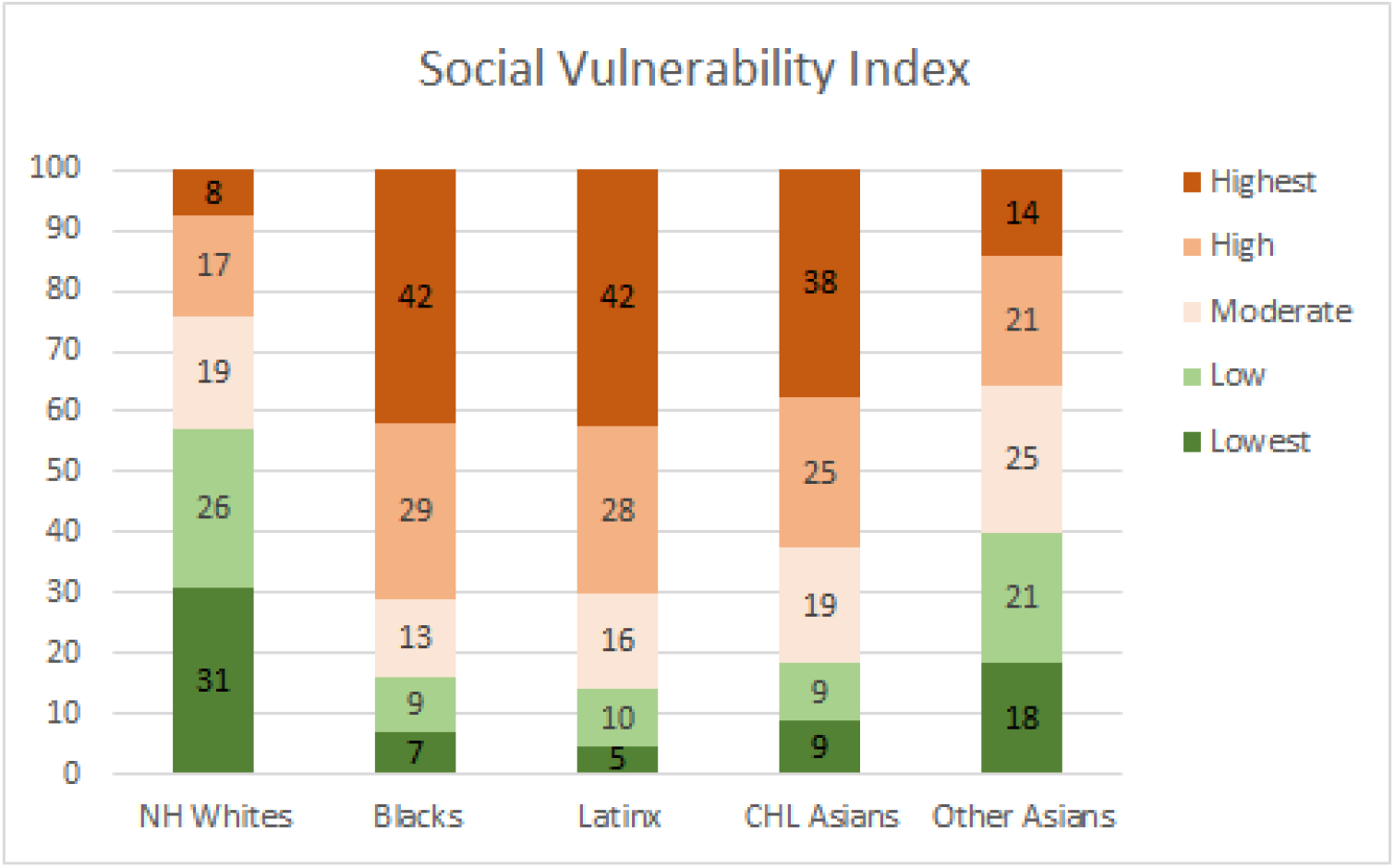
Ethnoracial Distribution by Neighborhood Social Vulnerability Index

### Vulnerability Levels By Racial Majority Groups

We further analyzed the four indicators of COVID-19 vulnerability by categorizing ZCTA’s into racial majorities and comparing their average vulnerability scores. Ethnoracial majority was assigned based on the racial and ethnic composition of each ZCTA. For instance, if a group makes up 50% or more of the population in the ZCTA then that group is assigned as being the majority ethnoracial group for that ZCTA. ZCTAs with no majority of one ethnoracial group are designated as “No Majority”.^v^ Figure 10 displays ZCTAs by their majority ethnoracial designation.

**Figure 10.**
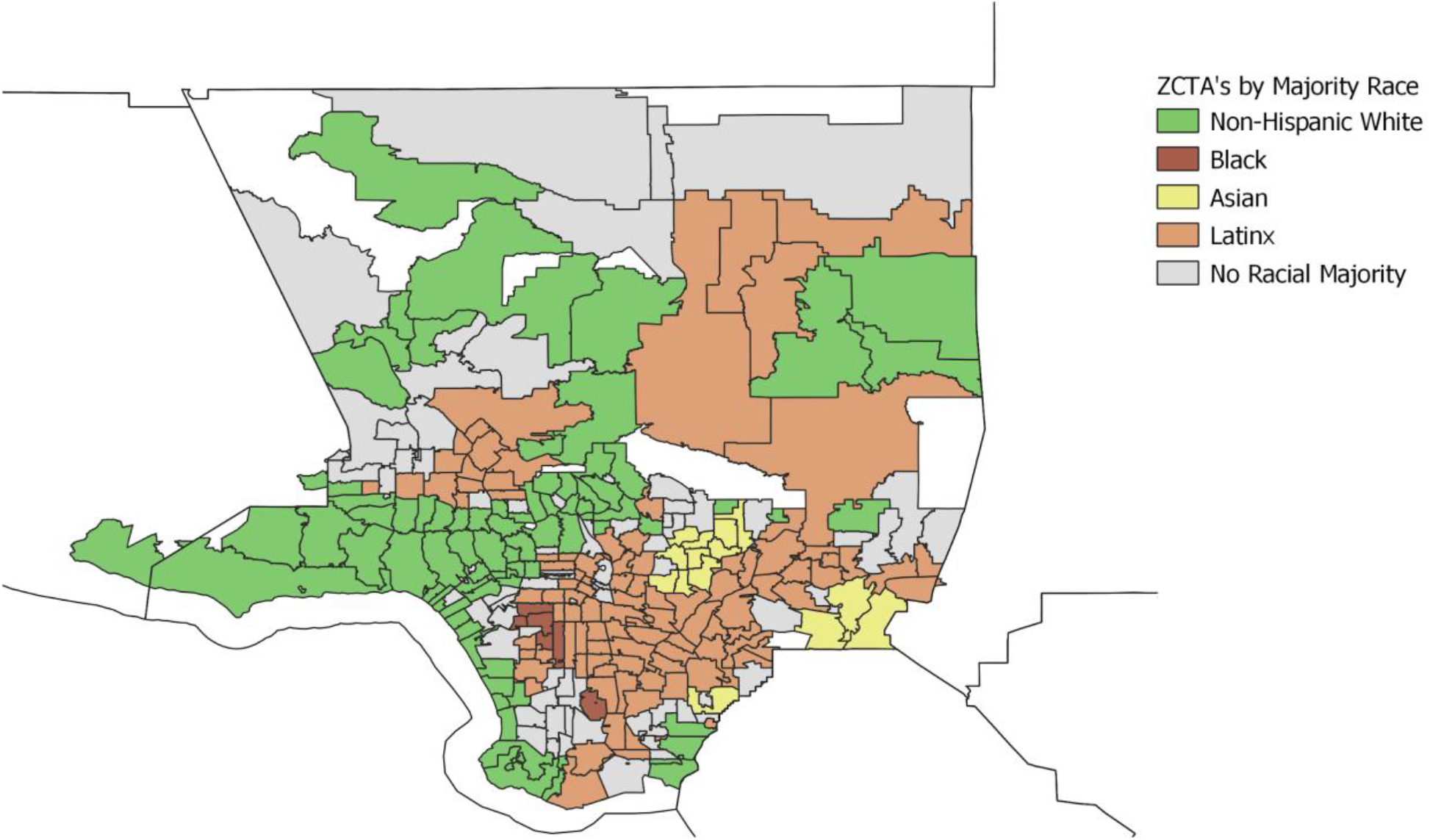
ZCTAs in Los Angeles County by Majority Ethnoracial Group. **Figure 10** identifies the majority ethnoracial group in each Zip Code Tabulation Area (ZCTA) as determined by the Bureau of Census.

We weighted each ZCTA by the area’s total population and then calculated the average vulnerability score for each neighborhood type. The higher the average values the more vulnerability. Across all four indicators, we see changes in the racial patterns of vulnerability. In Figure 11, majority Black and Latinx neighborhoods tend to have the highest pre-existing health vulnerability compared to other racial groups. In contrast, majority NH White neighborhoods tend to have the lowest level vulnerability. Additionally, as illustrated in Figure 12, neighborhoods with a majority Latinx population have the highest level of barriers to accessing healthcare, followed by neighborhoods with majority Asian populations. This is not surprising given that these two groups are largely immigrant populations and with language barriers. In contrast, majority NH white neighborhoods have the lowest level of barriers to accessing healthcare.

Figure 13 illustrates the distribution of built environment risk across racial groups. Majority Latinx neighborhoods tend to be more vulnerable to built environment risk compared to other groups. Interestingly enough, NH White, Asian, and Black majority neighborhoods are all within the low vulnerability range. Neighborhoods with a majority Black population have the lowest level of built environment risk. One probable explanation could be due to the location of these majority neighborhoods for Asians and Blacks, which in Los Angeles county, tend to be in less dense neighborhoods (out of the central city) with more single-family homes as in the case of Inglewood for African Americans and San Gabriel Valley for Asians.

**Figure 11.**
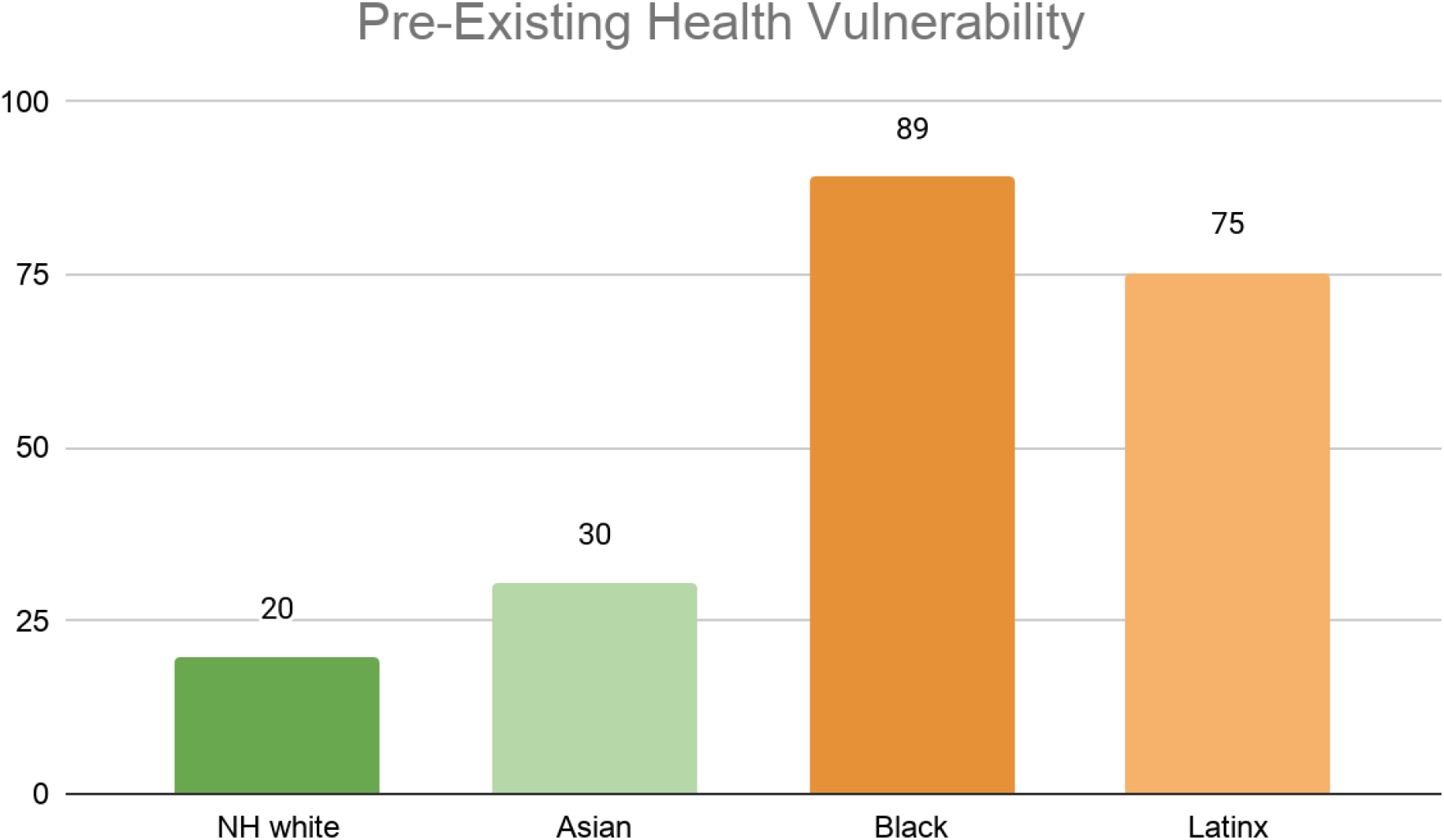
Average Pre-Existing Health Vulnerability Index by Los Angeles County Neighborhood’s Racial Majority

**Figure 12.**
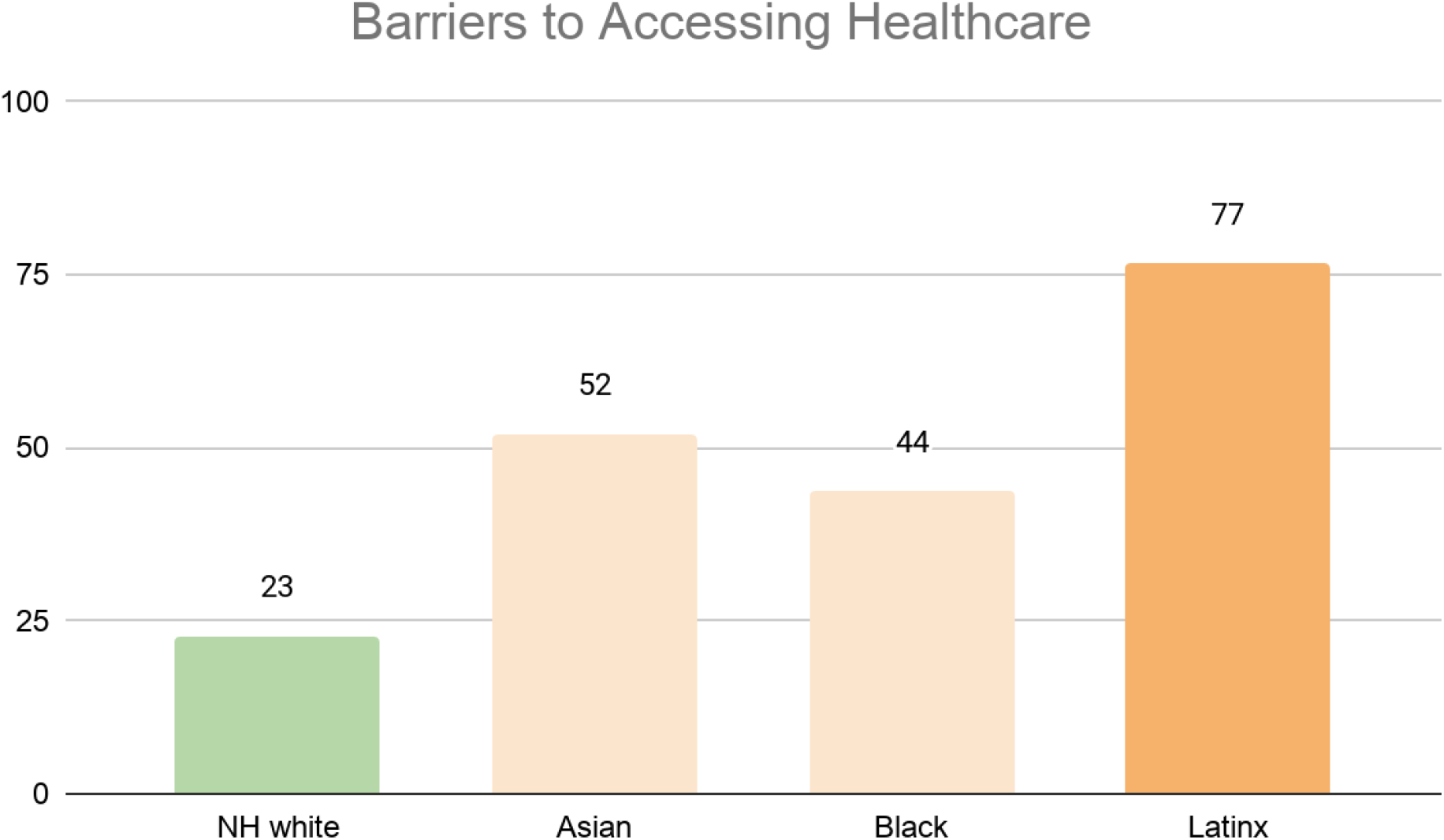
Average Barriers to Accessing Health Index by Los Angeles County’s Neighborhood’s Racial Majority

**Figure 13.**
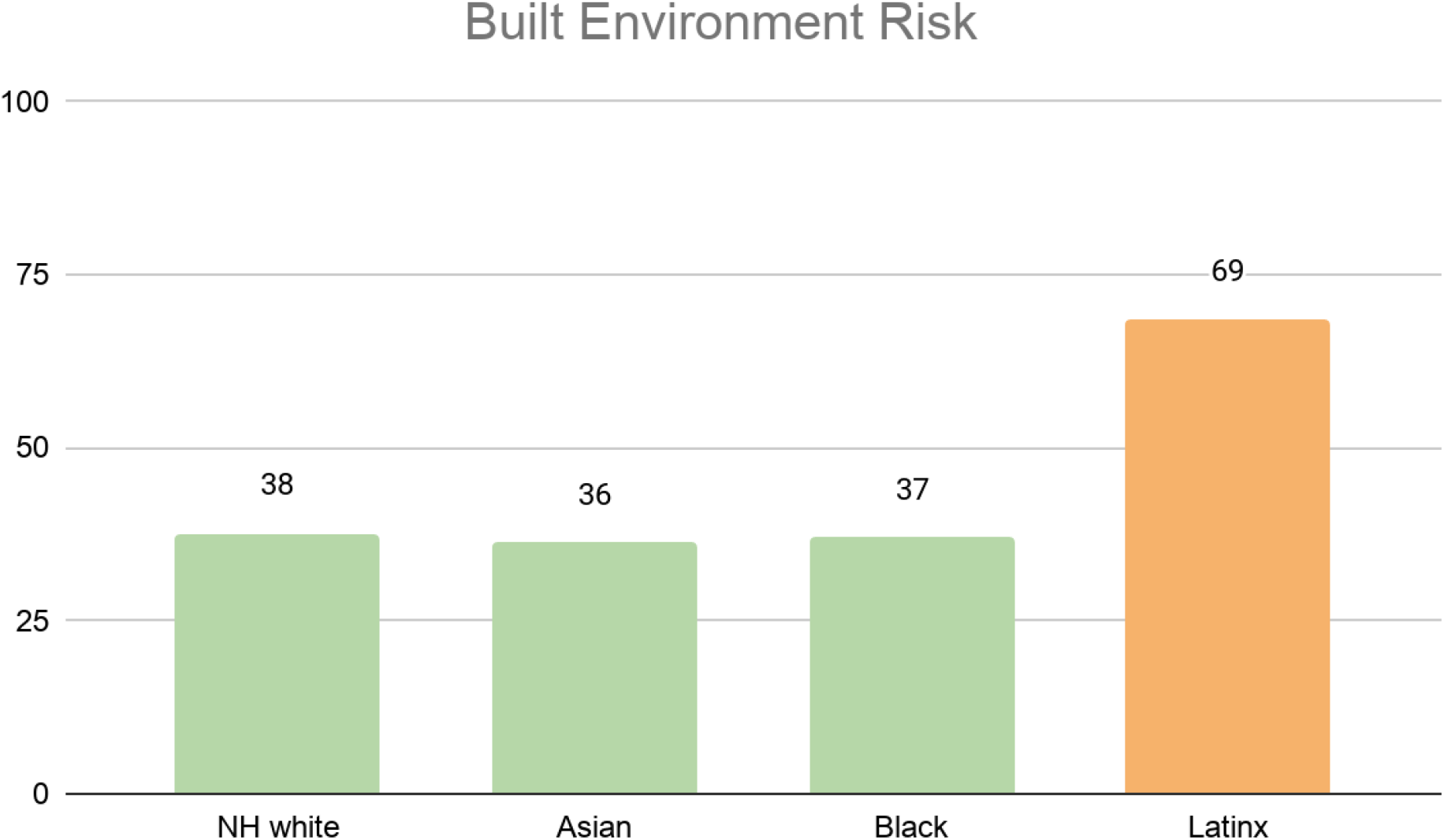
Average Built Environment Risk Index by Los Angeles Neighborhood’s Racial Majority

Finally, the Social Vulnerability Index, illustrated in Figure 14, shows that neighborhoods with majority Latinx and majority Black populations have the highest level of social vulnerability, while majority non-white neighborhoods have the lowest vulnerability.

**Figure 14.**
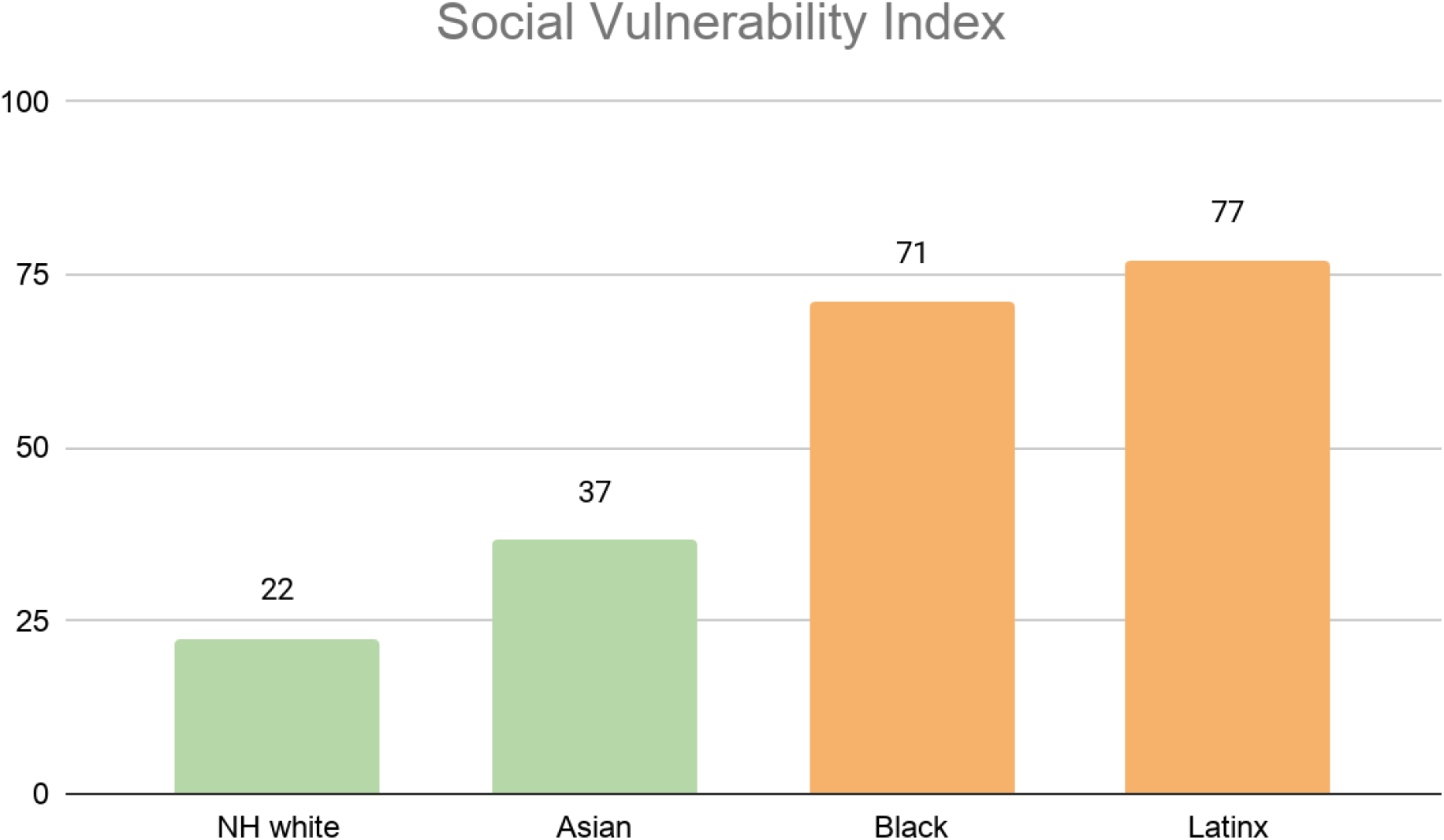
Average Social Vulnerability Index by Los Angeles County Neighborhood’s Racial Majority

### Neighborhood Socio-Demographic Characteristics by Levels of Medical Vulnerability

We also examine how each of the indicators correlate with neighborhood socio-demographic characteristics. The focus is on whether burdens are higher in disadvantaged communities (lower income, predominantly communities of color, and neighborhoods with a relatively large number of individuals with language barriers). It’s important to note that some of these socio-demographic variables analyzed are also embedded in some of the indices as underlying variables. We ranked neighborhoods from lowest to highest quintiles for each of the vulnerability indicators. The reported values represent the average (mean) of each variable for all the ZCTAs in each neighborhood group (quintiles).

The results in **Table 1**, illustrate the distribution of race, language barriers, and income across neighborhoods with different levels of pre-existing health vulnerability. Neighborhoods with the highest vulnerability disproportionately have a higher percentage of Latinx residents while neighborhoods in the lowest vulnerability have a higher percentage of non-hispanic white individuals. Additionally, neighborhoods with higher levels of vulnerability also disproportionately have more residents with language barriers (speaks English “less than well”) and have lower per capita income. For example, on average, the highest vulnerable neighborhoods have four times more residents with English language barrier than the lowest vulnerable neighborhoods. Additionally, the average per capita income in the highest vulnerable neighborhood is nearly 3.5 times less than the lowest vulnerable neighborhoods.

**Table 1.** Neighborhood Socio-Demographic Characteristics by Level of Pre-Existing Health Vulnerability

|  | Lowest | Low | Moderate | High | Highest |
| --- | --- | --- | --- | --- | --- |
| Distribution by Race |  |  |  |  |  |
| %NH white | 60% | 42% | 31% | 19% | 7% |
| %Black | 5% | 6% | 5% | 6% | 19% |
| %Latinx | 15% | 25% | 41% | 61% | 68% |
| %Asian | 16% | 24% | 20% | 12% | 4% |
| % Speaks English<br>“less than well” | 4% | 8% | 12% | 16% | 16% |
| Per Capita Income | \$67.4k | \$40.6k | \$31.9k | \$23.9k | \$19.6k |
| N (ZCTAs) | 51 | 52 | 52 | 53 | 52 |

Similar patterns are found in **Table 2, 3** and **4**, with the neighborhoods in the highest level of vulnerability for each of the indicators consisting of predominately Latinx and Black residents compared to neighborhoods in the lowest level of vulnerability. In contrast, the lowest vulnerable neighborhoods disproportionately have more non-Hispanic white residents. Furthermore, higher vulnerable neighborhoods across all indicators have a higher share of residents with English language barriers and lower income residents as opposed to the lower vulnerable neighborhoods.

**Table 2.** Neighborhood Socio-Demographic Characteristics by Level of Social Vulnerability

|  | Lowest | Low | Moderate | High | Highest |
| --- | --- | --- | --- | --- | --- |
| Distribution by Race |  |  |  |  |  |
| %NH white | 60% | 48% | 31% | 21% | 8% |
| %Black | 4% | 5% | 7% | 11% | 13% |
| %Latinx | 15% | 26% | 40% | 52% | 70% |
| %Asian | 16% | 18% | 19% | 14% | 9% |
| % Speaks English<br>“less than well” | 3% | 6% | 10% | 15% | 20% |
| Per Capita Income | \$63.9k | \$49.0 | \$33.3k | \$26.5 | \$18.6 |
| N (ZCTAs) | 55 | 56 | 55 | 56 | 55 |

**Table 3.** Neighborhood Socio-Demographic Characteristics by Level of Barriers to Accessing HealthCare

|  | Lowest | Low | Moderate | High | Highest |
| --- | --- | --- | --- | --- | --- |
| Distribution by Race |  |  |  |  |  |
| %NH white | 62% | 47% | 29% | 20% | 10% |
| %Black | 5% | 7% | 9% | 9% | 9% |
| %Latinx | 17% | 24% | 41% | 52% | 70% |
| %Asian | 12% | 18% | 18% | 18% | 10% |
| % Speaks English<br>“less than well” | 2% | 5% | 9% | 15% | 23% |
| Per Capita Income | \$66.0k | \$48.4k | \$31.7k | \$26.3k | \$19.0k |
| N (ZCTAs) | 55 | 56 | 55 | 56 | 55 |

**Table 4.** Neighborhood Socio-Demographic Characteristics by Built Environment Vulnerability

|  | Lowest | Low | Moderate | High | Highest |
| --- | --- | --- | --- | --- | --- |
| Distribution by Race |  |  |  |  |  |
| %NH white | 49% | 40% | 30% | 25% | 23% |
| %Black | 8% | 6% | 7% | 9% | 9% |
| %Latinx | 23% | 37% | 41% | 50% | 51% |
| %Asian | 16% | 14% | 19% | 13% | 14% |
| % Speaks English<br>“less than well” | 4% | 7% | 11% | 14% | 18% |
| Per Capita Income | \$51.4k | \$43.6k | \$36.3k | \$31.5k | \$28.5k |
| N (ZCTAs) | 55 | 56 | 55 | 56 | 55 |

## Discussion and Conclusion

This project developed four key indicators capturing different dimensions of COVID-19 related vulnerabilities. These indicators address multiple factors associated with increased risk of COVID-19 infections and mortality: pre-existing health conditions, barriers to accessing healthcare, built environment risk, and social vulnerability. We map each of the indicators to illustrate the different levels of vulnerability across Los Angeles County.

These four indicators share commonalities but are not identical. The analyses presented in the previous sections review similarities in terms of the most vulnerable neighborhoods. For instance, the most vulnerable neighborhoods across all indicators tend to be concentrated in areas within South Los Angeles and the San Fernando Valley. These places are marked by low income racial communities, while neighborhoods that showed up as being the least vulnerable are along the coastal regions of the county marked by less density, higher incomes and higher proportions of non-Hispanic Whites. In addition to mapping the four indicators, we presented a quantitative analysis of the socioeconomic and demographic profiles of the most and least vulnerable neighborhoods. Consistently across all indicators, the highest vulnerable are disproportionately people of color and low-income, relative to communities at the other end of the spectrum.

At the same time, there are variations across dimensions. The four indicators have low to high correlations (r values range from 0.38 for built environment risk and pre-existing health to 0.89 for SVI and barriers to accessing health care), indicating that each indicator is partially capturing unique elements of vulnerability. (See Appendix A for full correlation table). In other words, systemic inequality of vulnerability is complex, requiring multiple indices to fully understand the systematic pattern. This also means that addressing one’s vulnerability along one dimension does not automatically translate into addressing risks along the other dimensions. (For example, alternating colors and shades of colors in the above maps show that places in the northeastern section of Los Angeles County are not highly vulnerable to one type of risk but are highly vulnerable to another type of risk.)

The usefulness of the indicators varies with the types of policies and plans being developed to assist those most at risk from COVID-19’s direct and indirect impacts. For example, health planners would be more focused on pre-existing health conditions, and urban planners would be more focused on the built environment. It is the context of population health and the responsibilities of public health to address the health of its locale that data such as ours can serve as a roadmap. Public health is charged with decision-making of the distribution of health care resources, the multi-sectorial planning needed to address complex risk vulnerabilities and prevention efforts to stop or slow new infections. We provide a method for consideration of how resources to prevent new infections and associated morbidities should be provided to neighborhoods in the areas identified and prioritized by relevant indicators. We are providing information that combines medical needs with the social determinants of health outcomes which create risk and risk clusters in neighborhoods and in racial/ethnic populations.

We believe that the local data can enable public health agencies to better target scarce funds to improve the effectiveness of testing and monitoring of the disease. Local data can provide crucial knowledge and insights to social service providers, emergency agencies, and volunteers on where to direct their time and resources, such as where to set up distribution sites for food and other necessities. The ability to integrate these community factors to COVID-19 prevention plans is essential in determining where to secure hotel rooms for quarantine and social isolation. These informed and evidence-based actions are critical elements in slowing the spread of the disease. The data will continue to be valuable over the foreseeable future. Knowing where and who needs assistance allows responsible agencies to plan for the next wave of COVID-19 and for future pandemics to assist people dealing with housing, mental health, suicide and substance abuse issues.

While the information is useful to more efficiently allocate existing resources, we believe that more resources are needed. It is vital that state and local officials increase or shift funding in order to provide appropriate healthcare services and coverage for neighborhoods with high pre-existing health and social vulnerabilities. It is also necessary that state and local officials provide significantly more preventive resources and adequate health information to communities with high barriers to accessing healthcare. Furthermore, state officials and local policy makers should focus their efforts to physically transform neighborhoods with high built-environment risk and social vulnerabilities, to make these spaces safe for social distancing and quarantine efforts. Our data clearly show that communities with high built environment risk contain small living quarters, dense conditions and lack parks and greenery. We need community development that makes the neighborhood into healthy places.

It is our hope that our neighborhood and medical vulnerability indicators will be employed by local agencies to reduce and potentially halt new infections in areas of particularly high risk as a part of local efforts’ to move the county into conditions that can facilitate greater opening up of the city in order to support economic recovery. It is our hopes that these results can help guide Los Angeles County and even be used at the level of medical vulnerability indicators for the State of California as part of the science of openings that is coupled with equity considerations for some of the most vulnerable. These data also provide an equitable approach for the distribution of resources such as testing, vaccine priorities and provision of PPE and financial resources.

## Data Availability

Data used in this manuscript are available through the author.

## Contributor/Guarantor Information

The corresponding author attests that all listed authors meet authorship criteria and that no others meeting the criteria have been omitted. The study was conceptualized by PMO and VMM, indicators were developed by PMO, CP, NRG, VMM, analyses were conducted by PMO, CP, mapping was done by CP and NRG, first draft of the manuscript was written by PMO, CP, NRG and all authors edited and approved the final manuscript.

## Acknowledgments

We thank the UCLA’s BRITE Center on Science, Research & Policy (www.minoritydisparities.org) (MD 006932) for providing partial support for this research and analyses. This project builds on the UCLA Center for Neighborhood Knowledge’s (CNK) COVID-19 Equity Research Initiative, which includes studies examining how the negative economic impacts of COVID-19 are distributed across neighborhoods. We are grateful to the UCLA Latino Policy and Politics Initiative, UCLA Luskin Institute on Inequality and Democracy, UCLA Asian American Studies Center, UCLA American Indian Studies Center, UCLA Ziman Center for Real Estate and the University of California Office of the President (CBCRP Grant #R00RG2606) for their partnership and collaboration with the Initiative. These organizations supported previous work which serves as a partial foundation for the analytical work in this report. The authors alone are responsible for the content of this manuscript.

## Disclaimer

The views expressed herein are those of the authors and not necessarily those of the University of California, Los Angeles or the National Institute of Health as a whole. The authors alone are responsible for the content of this report.

## Footnotes

i The AskCHIS NE dataset also includes a separate variable on the health status of the elderly (ages 65 and older). We constructed two versions of the pre-existing health indicator, one that only includes ages 18-64 and an alternative that includes both ages 18-64 and 65 and up. The two indicators are highly correlated (r value of 0.9897). However, the version that includes the elderly had many more ZCTAs with no data, which is most likely a result of data suppression due to small sample size. The indicator including the elderly covers 251 ZCTAs, while the one that excludes the elderly covers 260 ZCTAs. Because the two are highly correlated and the indicator without the elderly covered more ZCTAs overall, we opted to use 18-64 in the final index.

ii We utilized the geographic crosswalk (ZCTAs to county) from the Missouri Data Center (2018 Geocorr) to identify ZCTAs in Los Angeles County. Only those ZCTAs that have at least an allocation score of 90% were included. The allocation factor indicates what proportion of the ZCTA, weighted by population, belongs in Los Angeles County (there are some ZCTAs that cross into neighboring counties).

iii For further details on the CDC’s SVI methodology see: https://svi.cdc.gov/Documents/Data/2018_SVI_Data/SVI2018Documentation.pdf

iv Los Angeles Times “Mapping L.A. Boundaries API”: http://boundaries.latimes.com/sets/. Retrieved on September 30, 2020

v Of the 284 ZCTAs utilized in this analysis, 84 are majority NH white, 6 majority Black, 101 majority Latinx, 15 majority Asian, and 78 “No Majority” ZCTAs.

