## Appendix A for "COVID-19 Medical Vulnerability Indicators: A Local Data Model for Equity in Public Health Decision-Making"

**Appendix A: Correlation of Vulnerability Indicators**

| **Prob > \|r\| under H0: Rho=0** | | | | |
| --- | --- | --- | --- | --- |
| **Number of Observations** | | | | |
|  | **Pre-Existing Condition** | **Social Vulnerability Index** | **Barriers to Accessing Healthcare** | **Built Environment Risk** |
| **Pre-Existing Condition** | 1 | 0.81828 | 0.73217 | 0.38435 |
|  |  | <.0001 | <.0001 | <.0001 |
|  | 260 | 259 | 259 | 259 |
| **Social Vulnerability Index** | 0.81828 | 1 | 0.89218 | 0.58566 |
|  | <.0001 |  | <.0001 | <.0001 |
|  | 259 | 277 | 277 | 277 |
| **Barriers to Accessing Healthcare** | 0.73217 | 0.89218 | 1 | 0.73708 |
|  | <.0001 | <.0001 |  | <.0001 |
|  | 259 | 277 | 277 | 277 |
| **Built Environment Risk** | 0.38435 | 0.58566 | 0.73708 | 1 |
|  | <.0001 | <.0001 | <.0001 |  |
|  | 259 | 277 | 277 | 277 |
